# Exploring the Interpretability of AI Decision Support Systems for Surgical Anatomy Recognition

**DOI:** 10.64898/2026.06.02.26354729

**Authors:** Danyal Z Khan, Tobias Adams, Anjana Wijekoon, Roxana Ramirez Herrera, Sophia Bano, Peter McCulloch, Danail Stoyanov, Matt Clarkson, Enrico Costanza, Ann Blandford, Hani J Marcus, CARES Evaluation Group

**Affiliations:** Queen Square Institute of Neurology, University College London, London, United Kingdom; National Hospital for Neurology & Neurosurgery, Queen Square, London, United Kingdom; Manchester University NHS Foundation Trust, Manchester, United Kingdom; UCL Hawkes Institute, University College London, London, United Kingdom; UCL Human Computer Interaction Centre, University College London, London, United Kingdom; Department of Surgical Sciences, University of Oxford, Oxford, United Kingdom

**Author notes:** equal contribution (Danyal Z Khan, Tobias Adams). **CARES Evaluation Group:** Mohammed M Adawi, Joao P Almeida, Jensen Ang, Anouk Borg, Alexandros Boukas, Aswin Chari, Lekhaj C Daggubati, Neil L Dorward, Samuel Emerson, Laurence J Glancz, Paul Gwammache, John G Hanrahan, Joachim Starup Hansen, Lauren Harris, Mark Hughes, Abhiney Jain, Adham M Khalafallah, Taufiq Khan, Muhammad S Khan, Ramez W Kirollos, Angelos G Kolias, Jacek Kunicki, Ruth Lau, Juan CF Miranda, Ramesh Nair, Nicola Newall, Amr Nimer, Fabio M Oddi, Piero A Oppido, Piotr Paździora, Hafiz MI Razzaq, Bosnjak Roman, Luis AF Salazar, Olabisi Sanusi, Thomas Santarius, Sebastian Senger, Jonathan Shapey, Nathan A Shlobin, Nicholas WM Thomas, Sebastian M Toescu, Georgios Tsermoulas, Juan F Villalonga, Adam Williams, Nina Yoh, Brett E Youngerman, Gabriel Zada. Affiliations at end of article. **Competing interests:** HJM is employed by and hold shares in Panda Surgical. DS holds shares in Panda Surgical, Odin Vision, and is employed by TouchSurgery, Medtronic. **Data availability:** Available upon reasonable request.

**Keywords:** Artificial intelligence, computer vision, decision support systems, surgery

## Abstract

Artificial intelligence (AI) decision support systems for surgery hold promise but face barriers to adoption, particularly around the interpretability of their outputs. We conducted an international cross-sectional survey of 47 neurosurgeons to evaluate perspectives on literature-derived explanation techniques for AI-generated anatomical segmentations, using endoscopic pituitary surgery as a high-risk exemplar. Participants ranked certainty scores, certainty maps, saliency maps, scene similarity scores, and nearest-neighbour illustrations, and rated them using a modified Explanation Satisfaction Scale alongside free-text feedback. Certainty-based techniques were consistently ranked and rated highest for interpretability — valued for aligning with surgical decision-making by conveying confidence (via scores) and anatomical boundaries (via maps). Saliency- and similarity-based methods were judged less clinically relevant and better suited to educational settings. Certainty-based explanations, therefore, appear most acceptable to surgeons for clinical integration of decision support systems, though their impact on AI acceptability, trust calibration, and performance requires prospective evaluation across surgical domains.

## Background

Artificial intelligence (AI) tools for surgical applications have demonstrated relatively slow uptake compared to other medical fields, particularly in decision support, despite their theoretical promise for improving safety, effectiveness and efficiency^1-3^. This is in part due to the complexity of deploying AI systems in the operating room environment. In order to analyse surgical procedures in detail, surgical computer vision (SCV), a rapidly expanding subtype of AI that analyses surgical video data, must be optimised for accurate real-time performance^4^. This is technologically challenging as surgical video data is high-dimensional, temporal, non-stationary with rapid viewpoint shifts, and often subject to occlusions and variable lighting^1, 2^. From a human factors perspective, surgery is already a high-stakes, cognitively demanding and dynamic environment, and so any technology introduced must be carefully integrated into the surgical workflow^5^.

Surgical anatomy recognition is a fundamental process in any surgical procedure, and has been targeted as a task which could be improved by AI assistance, synergising with existing tools that surgeons use for orientation and navigation (e.g. image-guidance)^6-9^. This leverages SCV technology, pioneered in histology and radiology for pathology detection, which has already been translated successfully into practice in those clinical domains^10^. Numerous AI decision support systems (AI-CDSS) to assist surgical anatomy recognition by segmenting key structures have been developed recently for a variety of surgeries, for example, laparoscopic cholecystectomy and endoscopic pituitary surgery, and have been shown to improve surgeon anatomy recognition and thus surgical safety^4, 9, 11, 12^. Pre-clinical AI-CDSS studies suggest that the interpretability of AI-CDSS output may strongly influence the appropriate uptake of AI recommendations^4, 12, 13^. Interpretable AI recommendations should facilitate clinician acceptability, appropriate trust calibration in AI tools, and maximise AI-CDSS effectiveness and safety (e.g. limiting potential over- or under-trust, or over- or under-reliance)^14^.

The interpretability of AI-CDSS outputs is influenced by AI model factors (e.g. accuracy, explainability), human factors (e.g. digital literacy) and AI-human interface factors (e.g. usability, cognitive intensity)^13^. The explainability of AI model output is a modifiable factor which can be optimised through targeted AI model and system interface design – improving user situational awareness of the AI tool’s behaviour and performance^15, 16^. Explanations include varying levels of information related to AI input (e.g. similarity of unseen data to seen data) or AI output (e.g. feature weights) and can be presented in many different formats, such as text, graphs, maps or decision tree hierarchies^17^. Ultimately, these explanation configurations should be evaluated from both the system designer’s perspective and the user’s perspective^18^. If explanation evaluations in surgical contexts do not demonstrate sufficient interpretability, surgeons may under- or over-trust the AI outputs, leading to inappropriate adoption and reliance. They must also present a careful balance of information at the right time, particularly for real-time surgical AI-CDSS^4, 13^.

The influence of explanations of AI output in surgical AI-CDSS interpretability, decision making and clinical impact is unknown. Its importance is underlined by recommendations from international guidelines in the clinical evaluation of AI systems, such as DECIDE-AI^19^. We sought to explore surgeons’ perspectives on the utility and interpretability of various literature-derived explanations of AI-CDSS outputs. We used endoscopic pituitary tumour surgery as an exemplar, where SCV-based AI-CDSS for anatomy recognition has already been developed, and where this anatomy recognition is amongst the highest risk of all surgeries owing to the tumour’s close proximity to life-sustaining neurovascular structures^9^. To our knowledge, this is the first study of its kind and has the potential to inform the design of AI-CDSS across any type of surgery.

## Methods

### Aim

The primary aim of this study was to evaluate different compositions of supplementary explanatory information for AI-generated anatomical segmentation outputs in endoscopic pituitary surgery. This evaluation is from the perspective of the end-user (surgeon) and will inform the design of future SCV tools to enhance intraoperative decision-making and surgical education.

### Study Design and Participants

A cross-sectional survey-based study administered electronically via the Qualtrics XM software program (Qualtrics, Seattle, WA) was adopted. Eligible respondents included neurosurgeons and neurosurgical trainees with a minimum of one year’s experience in performing the endoscopic transsphenoidal pituitary surgical approach. The study was advertised through national neurosurgical organisations (predominantly UK-based), trainee networks (predominantly UK-based), and social media channels (internationally, via X and LinkedIn).

### Ethical considerations

All methods were carried out in accordance with the principles of the Declaration of Helsinki and were approved by the University College London Ethics Committee (UCLIC-2024_009). Participation was voluntary, and informed consent was obtained from all participants. Participants were offered collaborative authorship as part of the “CARES Evaluation Group” to recognise their contribution.

### Survey Content and Structure

Baseline characteristics were collected – including demographics (age, biological sex); clinical experience in performing endoscopic transsphenoidal pituitary surgery; and frequency of respondent interaction with AI tools in their clinical or academic practice.

Participants then received a brief explanation of the underlying AI model featured in subsequent sections, representing the minimum level of information necessary to understand the tool’s function. Details of this tool have been described elsewhere^9^. Participants were then shown the same still frame from the sellar phase of pituitary surgery with an illustrative AI overlay incorporating one of five different supplementary explanatory techniques: certainty scores, certainty maps, saliency maps, scene similarity scores and nearest neighbours illustration (Figure 1). The order in which techniques were presented was randomised for each participant. The choice of explanatory techniques was inspired by the wider literature and the SAFE-AI guidelines, which organise explainable AI techniques into Level 1 or “Perception” (e.g. certainty metrics), Level 2 “Comprehension” (e.g. saliency maps) and Level 3 “Projection” (scene similarity techniques)^16, 20-25^. After reviewing each technique individually, participants completed a modified Explanation Satisfaction Scale (ESS) to provide contextualised Likert-based ratings of explanatory technique understandability, satisfaction, detail sufficiency, usefulness and accuracy^18^. This was accompanied by an open question on each technique’s utility for clinical or educational applications, and wider general feedback. Finally, participants were presented all five explanatory techniques together, along with just the AI overlay, without supplementary explanatory information, and asked to rank by preference.

**Figure 1.**
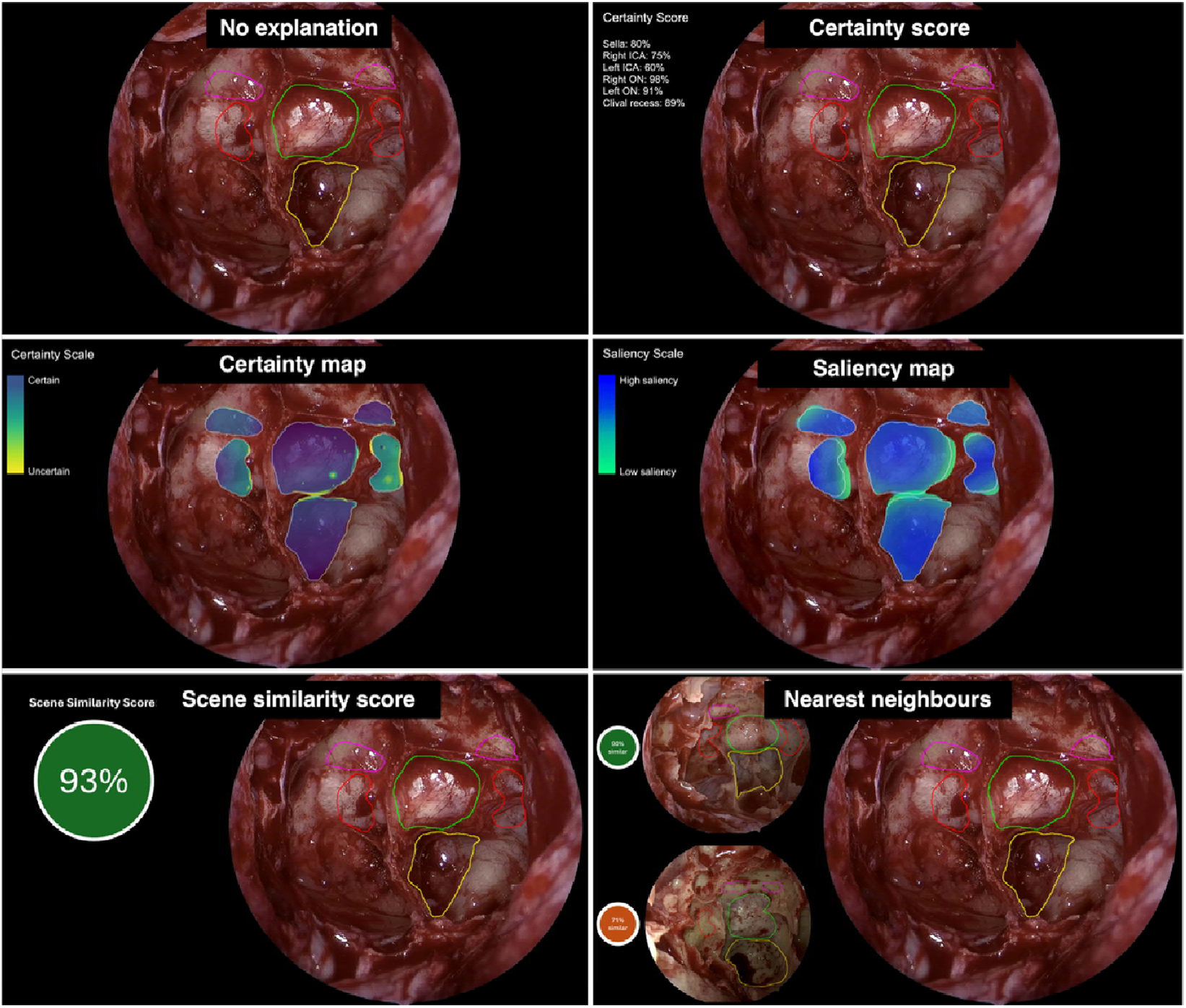
Overview of explanatory techniques presented to surgeons during the study. The Certainty Score was described as a quantification of the AI model’s certainty in identifying the area of each anatomical structure, representing the average pixel-wise certainty across the entire structure as a percentage^20^. The Certainty Map was described as a display of the AI model’s certainty in identifying the area for each anatomical structure, with dark blue indicating strong certainty, while yellow-green indicated uncertainty^21^. The Saliency map was described as the regions in the image that the AI model considered most important for identifying anatomical structure, with dark blue indicating high saliency (where the model focuses most of its attention), while green-turquoise represented lower saliency^22, 23^. The Scene Similarity Score was described as a numeric representation of how closely the current surgical scene matched the examples the AI model was trained on, with a higher percentage suggesting the model is working within its “comfort zone,” making it more likely that its predictions are accurate^24, 25^. The Nearest Neighbours technique was described as an illustration of how similar the current image is to examples the AI was trained on and provides two “nearest neighbour” images for comparison, along with their scene similarity scores relative to the primary surgical view^24, 25^.

### Data Analysis

Descriptive summary statistics were produced for baseline characteristics, ranking and ESS scores. For ranking data, a weighted average ranking was calculated by dividing the weighted sum by the total number of rankings per explanatory technique. The weighted sum for each structure was calculated as the sum of the rank frequency at each position (1-6), multiplied by the corresponding rank position value (1^st^ = 1, 2^nd^ =2, ). Free-text responses were thematically analysed using an inductive approach to identify the 3 most prominent feedback domains per explanatory technique.

Comparative statistics were performed using Excel (V16.1, Microsoft, USA) and R (V4.4.3, R Studio). Differences in technique rankings and ESS scores were assessed using the non-parametric Friedman test for repeated measures. For each analysis, the Friedman χ^2^ statistic, degrees of freedom (DoF = k–1), and p-values were reported. Effect size was quantified using Kendall’s coefficient of concordance (W), which ranges from 0 (no agreement) to 1 (perfect agreement). Post hoc pairwise Wilcoxon signed-rank tests were conducted with Holm correction for multiple testing, with effect sizes reported using Cohen’s R (ranging from 0, indicating no agreement, to 1, indicating perfect agreement). Spearman’s correlation coefficient was calculated to assess the relationship between mean ESS scores and weighted average rank to assess the consistency of surgeon feedback.

## Results

### Sample characteristics

An international cohort of 47 neurosurgeons completed the study survey (Table 1). The majority of participants practised in the UK (n=22, 46.8%) or USA (n=11, 23.4%). Other countries of practice represented were: Italy (n=2, 4.3%), Pakistan (n=2, 4.3%), Singapore (n=2, 4.3%), Poland (n=2, 4.3%), Egypt (n=1, 2.1%), Chile (n=1, 2.1%), Germany (n=1, 2.1%), Argentina (n=1, 2.1%), Slovenia (n=1, 2.1%) and Spain (n=1, 2.1%). The cohort was diverse in terms of age, surgical experience and experience with AI-assisted technologies (Table 1). Of note, most participants were attending level (68.1 %), and the majority used AI for clinical or academic tasks at least once a month (70.2%).

**Table 1:** Participant Demographics and AI Experience. †Participants could select multiple options; therefore, percentages are calculated based on the total number of selections and may not sum to 100%.

| Characteristic | Category | Count (N=47) | Percentage (%) |
| --- | --- | --- | --- |
| Age Distribution | 25-34 years old | 11 | 23.4 |
|  | 35-44 years old | 24 | 51.1 |
|  | 45-54 years old | 5 | 10.6 |
|  | 55-64 years old | 5 | 10.6 |
|  | 65+ years old | 2 | 4.3 |
| Biological Sex | Male | 40 | 85.1 |
|  | Female | 6 | 12.8 |
|  | Prefer not to say | 1 | 2.1 |
| Professional Role | Neurosurgery Consultant or Attending | 32 | 68.1 |
|  | Neurosurgery Registrar or Resident | 13 | 27.7 |
|  | Neurosurgery Senior Fellow (post-training) | 2 | 4.3 |
| Years of Experience Performing Endoscopic Pituitary Surgery | 1 to 5 years | 20 | 42.6 |
|  | 5 to 10 years | 15 | 31.9 |
|  | 10 to 20 years | 8 | 17.0 |
|  | Over 20 years | 4 | 8.5 |
| AI Usage Areas† | Academic research | 28 | 31.1 |
|  | Administrative tasks | 18 | 20.0 |
|  | Clinical decision-making | 12 | 13.3 |
|  | Disease diagnosis | 6 | 6.7 |
|  | Medical imaging interpretation | 7 | 7.8 |
|  | Robot-assisted surgery | 6 | 6.7 |
|  | None | 12 | 13.3 |
| AI Interaction Freq. | Every day | 11 | 23.4 |
|  | Every week | 14 | 29.8 |
|  | At least once per month | 8 | 17.0 |
|  | At least once per year | 4 | 8.5 |
|  | Never | 10 | 21.3 |

### Comparative Analysis of Explanatory Techniques

Certainty-based explanatory techniques – spatial maps and numeric per-structure summary scores – emerged as the top-ranked and top ESS scored options by surgeons (Tables 2-4, Figure 2).

**Table 2:** Summary of modified Explanation Satisfaction Score results for each technique.

| Technique | Certainty map | Certainty score | Scene similarity score | Nearest neighbours | Saliency map |
| --- | --- | --- | --- | --- | --- |
| This explanatory technique is **satisfying**. | 4.32 | 4.13 | 3.96 | 3.70 | 3.91 |
| This explanatory technique shows me how **accurate** the AI prediction is. | 4.32 | 4.38 | 4.11 | 3.64 | 3.53 |
| This explanatory technique has sufficient **detail**. | 4.30 | 4.19 | 3.51 | 3.74 | 3.64 |
| Using this explanatory technique, I **understand** how the AI works. | 4.00 | 3.85 | 3.70 | 3.83 | 3.62 |
| This explanatory technique is **useful** to my surgical or educational goals. | 3.96 | 3.85 | 3.70 | 3.49 | 3.38 |
| Mean Score | 4.18 | 4.08 | 3.80 | 3.68 | 3.62 |

**Figure 2.**
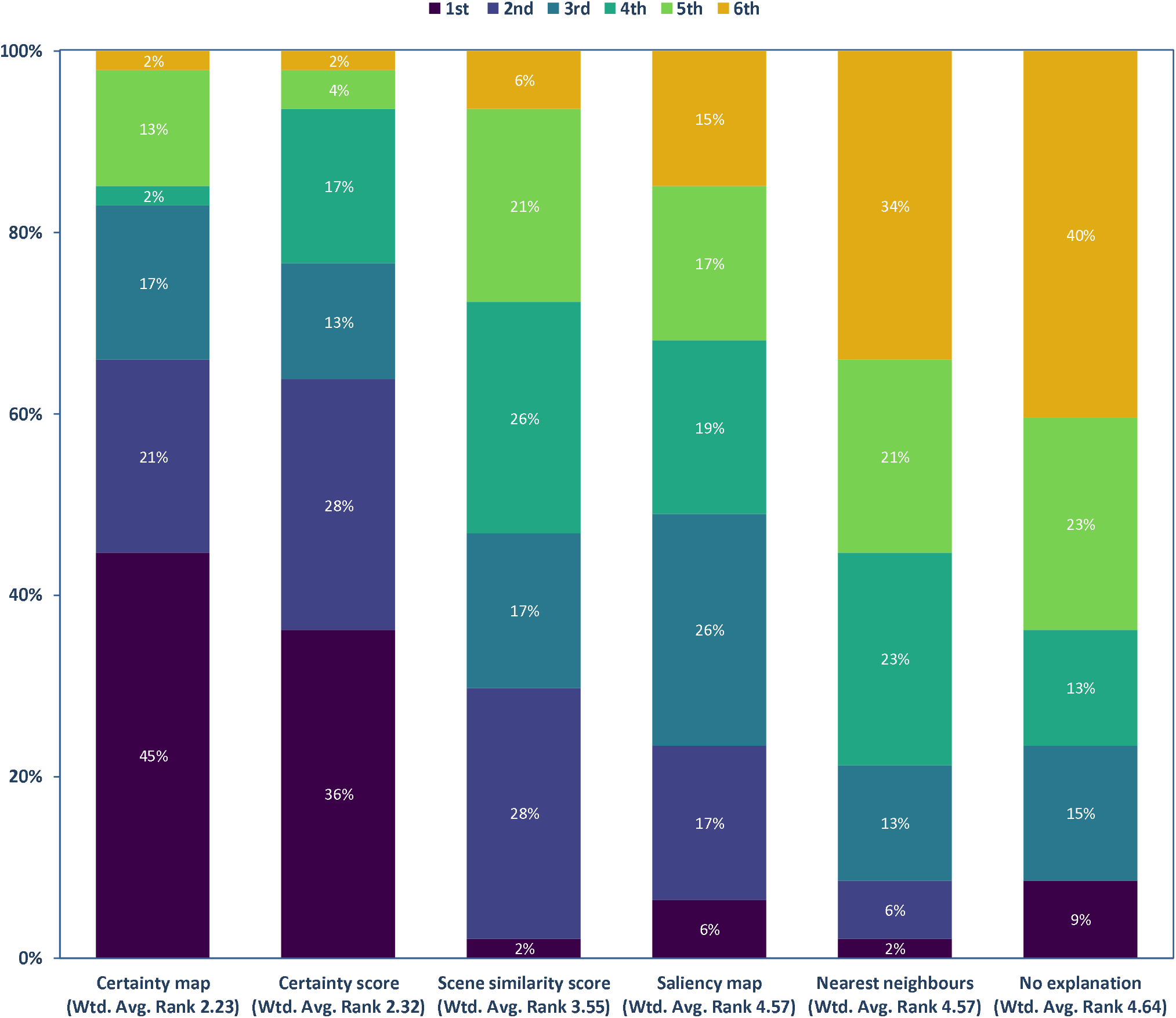
Stacked bar chart display of ranking frequency per structure. Wtd. Avg. Rank = Weighted Average Rank.

Firstly, there was a significant difference in ESS scores between techniques (Friedman test: χ^2^=32.1, p<0.001, W=0.17). Certainty map and certainty score were rated significantly higher than saliency maps, scene similarity scores, and nearest neighbours on post-hoc pairwise Wilcoxon signed-rank tests (Table 3). There was no significant difference between the certainty map and certainty score (Table 3; p=0.50, r=0.22).

**Table 3:** Pairwise Wilcoxon Signed-Rank Test Results for ESS Scores, with p-values and corresponding effect size via Cohen’s r. Significant comparisons (p < 0.05) are marked with an asterisk (*). Diagonal cells (—) indicate comparisons of a technique with itself. ESS = explanation satisfaction scale.

| Technique | Certainty map | Certainty score | Scene similarity score | Nearest neighbours | Saliency map |
| --- | --- | --- | --- | --- | --- |
| Certainty map | — | 0.503 (r=0.22) | 0.038* (r=0.40) | 0.006* (r=0.49) | <0.001* (r=0.64) |
| Certainty score | 0.503 (r=0.22) | — | 0.049* (r=0.38) | 0.006* (r=0.49) | 0.002* (r=0.54) |
| Scene similarity score | 0.038* (r=0.40) | 0.049* (r=0.38) | — | 0.567 (r=0.16) | 0.503 (r=0.22) |
| Nearest neighbours | 0.006* (r=0.49) | 0.006* (r=0.49) | 0.567 (r=0.16) | — | 0.617 (r=0.07) |
| Saliency map | <0.001* (r=0.64) | 0.002* (r=0.54) | 0.503 (r=0.22) | 0.617 (r=0.07) | — |

Regarding ranking, there was a significant difference in rankings between techniques (Friedman test: χ^2^=73.6, p<0.001, W=0.31). Certainty map and certainty score were ranked significantly higher than saliency maps, scene similarity scores, and nearest neighbours on post-hoc pairwise Wilcoxon signed-rank tests (Table 4). Again, there was no significant difference between certainty map and certainty score (Table 4; p=1.0, r=0.04).

**Table 4:** Pairwise Wilcoxon Signed-Rank Test Results for Rankings, with p-values and corresponding effect size via Cohen’s r. Significant comparisons (p < 0.05) are marked with an asterisk (*). Diagonal cells (—) indicate comparisons of a technique with itself.

| Technique | Certainty map | Certainty score | Scene similarity score | Nearest neighbours | Saliency map |
| --- | --- | --- | --- | --- | --- |
| Certainty map | — | 1.000 (r=0.04) | 0.005* (r=0.49) | <0.001* (r=0.69) | <0.001* (r=0.74) |
| Certainty score | 1.000 (r=0.04) | — | 0.002* (r=0.54) | 0.004* (r=0.52) | <0.001* (r=0.87) |
| Scene similarity score | 0.005* (r=0.49) | 0.002* (r=0.54) | — | 1.000 (r=0.04) | 0.005* (r=0.49) |
| Nearest neighbours | <0.001* (r=0.69) | 0.004* (r=0.52) | 1.000 (r=0.04) | — | 0.073 (r=0.36) |
| Saliency map | <0.001* (r=0.74) | <0.001* (r=0.87) | 0.005* (r=0.49) | 0.073 (r=0.36) | — |
| No explanation | <0.001* (r=0.81) | <0.001* (r=0.81) | 0.005* (r=0.50) | 0.073 (r=0.34) | 1.000 (r=0.05) |

Spearman’s test reveals a strong, negative correlation between the mean ESS Scores and the weighted average ranks (correlation coefficient -0.90; p=0.037), suggesting high concordance between ESS scores and rankings.

### Certainty Map: Quantitative & Qualitative Feedback

The certainty map explanatory technique had the highest average rank, the highest mean ESS score and the highest mean score in all domains except in the conveying accuracy domain (Table 2, Figure 2). As a spatial display of the certainty score, it appeared to improve granularity, comprehension and perceived utility, but sacrificed simplicity in conveying AI accuracy (Tables 2&3). This was reflected in the thematic analysis which highlighted the ***surgical utility of visual-spatial feedback*** for these complex 3d anatomical structures – particularly with respect to recognizing the limitations of the AI overlays (e.g., “indicates where I may need to pay more attention… to determine anatomic boundaries”, “which areas may require further adjuncts”, and “areas of uncertainty are helpful additions, as they could help guide decision making, and inform the degree to which the AI should be trusted”). This translated into ***the potential for intuitive improvements to training and surgical performance***– this was particularly the case for offline educational applications (“very helpful for teaching as helps with anatomical education”). However, its application in real time during surgery split opinions – with some surgeons for (e.g. “surgically this is the most useful…especially the medial border of the ICA, where certainty happens to be low so great caution needed in following this too closely”) and some against (“I am not sure about its utility for decision support” and “I don’t think heat maps would be particularly useful during live surgery”). This is likely in part due to the overlay design and its ***potential for information overload***, especially in real time, with this technique (“it seems a bit cluttered and obstructive of the surgeon’s view” and “too busy for my taste”)

### Certainty Score: Quantitative & Qualitative Feedback

The certainty score, presented on a per-structure basis (Fig. 1), had the second-highest average rank and mean ESS score (Table 2, Figure 2). It had the highest score in the conveying accuracy ESS subdomain – reflecting its simplicity and specificity as an explanatory technique. On analysis of the qualitative feedback on the impact of certainty scores on the clinical utility (e.g. teaching or decision support) of the AI overlay, key themes revolved around the pros and cons of a numeric value. Many respondents appreciated the ***value of simplistic quantification***, allowing “the surgeon to decide quickly” using “concrete” information which would be “neat to display in real-time”, whilst potentially reducing cognitive overload (“provides rapid support, especially in case of mental fatigue of the surgeon”). However, ***uncertainty in utility for real-world decision-making*** was expressed, emphasising the need for better definitions of what numeric value is meaningful (“what cut off is used for clinical decision making is uncertain - is 75% certain good enough to allow you to drill next to carotid?” and “it might be prone to an anchoring effect…I might think 80 is ok if other scores are in the 60s, or I might think it’s unacceptable if all the others are in the 90s”). Furthermore, ***lack of granularity and nuance*** emerged as a recurring concern, particularly for users who voluntarily described themselves as “visual learners”. Specifically, the lack of spatial information was cited repeatedly (“does not yield information regarding which areas may be less certain”). Furthermore, some users felt visual displays were more cognitively demanding, especially if displayed separately (“may slow down application of AI in a surgical setting as based on reading numbers…easier to rely on colour maps” and “prefer more visual indicators vs numbers as may be a bit more challenging to interpret fully intra-op if lots of info on screen”). This issue seemed to in part be due to the certainty score’s numeric nature but also in its presentation, separate from the overlays in a legend (“difficult to look away from the camera to read the numbers” and “Separating data presentation increases cognitive load… I find it strenuous to interpret the confidence of annotations with this format”).

### Scene Similarity Score: Quantitative & Qualitative Feedback

After the certainty-based techniques (map and score), the scene similarity score technique had the next highest average rank and mean ESS score (Table 2, Figure 2). On thematic analysis, surgeons found it ***over generalized and lacking specificity*** (e.g., “a single score for all structures within the scene generalizes too much”) and ***challenging to interpret*** (e.g., “To use it on its own would be too much of a black-box”, “does not explain what was similar/different between this image and the examples that the model was trained”, and “what the model perceives to be similar is NOT necessarily actually similar, this does not increase my confidence in the classification at all”). Like with the certainty score, the ***uncertain actionability*** was expressed (e.g., “clinically meaningful threshold would be good to include” and “% for each anatomical structure would be more helpful”).

### Nearest Neighbours: Quantitative & Qualitative Feedback

This technique ranked last in average rank and second last in mean ESS scores (Table 2, Figure 2). By leveraging examples of similar cases, it performed relatively well in conveying an understanding of how the AI worked (Table 2). Core themes emerging from surgeon feedback on its utility were concerns about ***information overload potential*** (e.g. “too confusing”, “too much information in real-time” and “showing three images simultaneously would be confusing”) and the ***questionable clinical relevance of similarity*** (“even minor variations with high scene similarity will have significant surgical relevance”). Nevertheless, there was an acknowledgement of its potential role for ***early education of anatomical variations*** aimed at less experienced members of the clinical team.

### Saliency Map: Quantitative & Qualitative Feedback

Saliency Map was the least effective technique overall according to the mean ESS score, and ranked second last in rankings (Table 2, Figure 2). It scored particularly poorly on its usefulness to users’ goals (Table 2), suggesting that while visually engaging, simply highlighting where an AI is “looking” is insufficient for user satisfaction and trust. Thematic analysis reflected this ***ambiguous clinical relevance*** (e.g., “whilst this helps understand how the AI works and generates its suggestions, I don’t see how this particular visualisation is helpful in training or for decision support” and “you know it is looking there but not whether it’s confident about its predictions”). Furthermore, when compared to certainty maps, surgeons felt saliency maps demonstrated a ***mismatch between AI focus and clinical priorities*** (e.g. “focuses more on the most obvious anatomical delineations and less on the neighbouring or transitional areas, which are the most dangerous from a surgical point of view”).

### Wider feedback

Thematic analysis of optional wider feedback distilled 3 key themes. Firstly, an emphasis on ***user customisation*** in designing such AI systems to allow for differences in preference and clinical workflow – specifically, the option to toggle between numeric and visual displays (e.g. the certainty score and certainty map). Secondly, consideration for ***hybrid techniques*** was requested to synergise the benefits: for example, combining the certainty score and certainty map techniques. Finally, numeric values were considered more feasible for use as real-time intra-operative decision support, whilst visual displays were felt to be particularly useful for post-operative training and education.

## Discussion

### Principal Findings

This study provides novel insights into surgeon perspectives on the design of user-centred surgical computer vision-based AI-CDSS. Using an existing anatomy recognition assistance computer vision AI model developed for endoscopic pituitary surgery as an exemplar, we explored the impact of a variety of literature-derived techniques which seek to make the decisions of otherwise “black-box” AI systems more interpretable, which in turn may impact trust, safety and translational impact. To our knowledge, this is the first study of its kind in surgical computer vision, addressing the unique challenges surgery presents when compared to imaging, histopathology or diagnostic endoscopy computer vision applications.

We found that metrics that are based on the concept of certainty, that is, how certain an AI model was in its predictions (i.e. epistemic uncertainty), aligned most closely with surgeons’ perspectives and cognitive workflow. Quantifying uncertainty is a core tenet of clinical decision-making, particularly in the context of using decision support systems, as it helps clinicians critically appraise AI assistants and judge their reliability and trustworthiness^26-29^. The Certainty Map, a visual-spatial representation of this, received the highest overall ESS score and was the most frequently top-ranked technique. This preference was driven by its perceived utility in providing intuitive feedback on anatomical boundaries between structures and areas of uncertainty where additional adjuncts may need to be utilised (e.g., Doppler ultrasound or image-guidance). The existing surgical thought processes and workflows were cited as a crucial consideration for enhancing safety and informing decision-making. The Certainty Score, which provides a summary numeric value for each identified anatomical structure, also performed well, scoring highest in the “conveying accuracy” ESS domain. Its simplicity was seen as a key advantage for rapid interpretation, particularly in a cognitively demanding intra-operative environment. Hybrid numeric and visual designs or the ability to toggle between them were endorsed by multiple participants.

Conversely, techniques that focused on the internal workings of the AI without direct clinical relevance - namely the Saliency map, Scene similarity score, and Nearest neighbours - were ranked significantly lower. Of these, scene similarity scores performed best in the conveying accuracy ESS subdomain, potentially reflecting its interpretation simplicity as a numeric value, akin to the certainty score. These techniques were suggested to have greater potential for educational purposes, particularly for junior trainees learning to identify anatomical variations, but were deemed unsuitable for real-time intraoperative use. Practically, out-of-distribution data and/or domain shift (i.e. the operative scene analysed is significantly different to the data the AI model was trained on) can be inferred from certainty estimates, and therefore scene similarity and nearest neighbour type displays may be functionally redundant^29^.

### Findings in the Context of the Literature

Our findings resonate with a growing body of literature on the critical role of techniques to improve AI interpretability and the wider human-computer interaction domain in bridging the translational gap of surgical AI systems – improving their usability and safety, and fostering clinician trust and collaboration. This is particularly relevant for deep learning methods, which are inherently “black box” in terms of their decision-making processes^30^. The preference for certainty-based explanations, such as certainty maps and scores, aligns with recent work highlighting that transparent communication of an AI model’s limitations is a cornerstone of responsible clinical deployment^31^. A recent review on explainable AI for medical imaging suggested that providing uncertainty estimations can improve the trust calibration of clinicians, preventing both over- and under-reliance^32^. The Certainty map, in particular, by translating abstract uncertainty into an intuitive visual format, provides a level of comprehension that surpasses simple numeric metrics, reflecting its high ranking and ESS scores in our study. This is a critical distinction, as some studies suggest that oversimplification of AI output can lead to an “anchoring effect” where clinicians may inappropriately trust a single high-certainty number without understanding the underlying spatial or contextual nuances, a concern explicitly raised by participants in our study^29^. Possible strategies to mitigate anchoring bias further could include encouraging users to make initial decisions themselves before utilising AI assessment, which could be enabled in this context by allowing users to toggle the AI overlay on or off^33^. However, whilst conceptually appealing, these methods need further study as to how they should be safely integrated into decision-making - for example, certainty thresholds aid interpretation of what is acceptably reliable; how to monitor whether certainty metrics are well calibrated to model accuracy and thus how to mitigate against the dangers of overconfidence (i.e. incorrect AI prediction, but with high certainty estimates); and whether the use of certainty metrics actually improves surgeon acceptability, performance and trust calibration^4, 27, 29^. Furthermore, many techniques that are used to estimate certainty or uncertainty in neural networks (e.g. Bayesian networks) are computationally expensive and may hamper real-time performance, although the rate of advancements in computing (e.g. specifically edge computing) suggests this is a time-limited issue^29^.

The challenges observed with saliency maps are also well-documented. While a popular technique in technical AI literature, saliency has been criticised in medical contexts for not providing a sufficient causal link between the model’s focus and the clinical task at hand^34^. Clinicians need to know not just where the model is looking, but why those areas are important and what the confidence level is for the resulting prediction. This study confirms that for a high-stakes task like identifying critical neurovascular structures, an explanation that lacks this crucial contextual and confidence information is of limited value.

### Strengths and Limitations

This international study was built on a foundation of existing evidence – specifically an AI system with demonstrated surgeon benefit for use as an exemplar case study, and optimised AI overlay designs, such that focus on wider design considerations, beyond the AI overlay or clinical use case, could be focused on^21, 35^. Other strengths include the literature-derived nature of the selected explanatory techniques and the combination of validated quantitative and insightful qualitative feedback attained, which we believe can be extrapolated to other surgical computer vision applications. Limitations include the UK- and US-centric sample recruited despite best efforts to disseminate the opportunity widely – likely reflecting the natural networks of the author group – and reducing its representativeness of the global surgical community. Additionally, the analysis was performed on still, anonymised images rather than in a real-time, intra-operative environment. The preferences observed may change with repetitive usage of the tool over time and when faced with the dynamic challenges of live surgery (e.g., bleeding, rapid viewpoint shifts and increased cognitive overload). Furthermore, respondents may have rated explanatory techniques as satisfying or dissatisfying when in fact their understanding of the process being explained was piecemeal or flawed^18^. Finally, the actual impact of the addition of these explanatory techniques on educational or surgical outcomes and on trust needs further study via prospective trials.

## Conclusions

This international study of surgeon perspectives on techniques to improve the interpretability of otherwise “black box” surgical computer vision AI systems showed that methods focused on conveying the certainty of AI system predictions appear superior. In high-risk applications, such as critical anatomy recognition assistance, conveying certainty numerically and visually (as hybrid displays or as toggled options) aligns most closely with surgical workflow and demonstrates potential for fostering well-calibrated trust in these systems. Further studies are needed to explore the impact of integrating surgeon-centred interpretability-improving techniques in AI-CDSS on clinical performance and trust calibration.

## Data Availability

Available upon reasonable request.

## Funding

This work was funded by the EPSRC (EP/Y01958X/1 and EP/W00805X/1). It was supported more generally by the UCL Hawkes Institute, formerly the Wellcome/EPSRC Centre for Interventional and Surgical Sciences (WEISS) (203145/Z/16/Z). DZK is supported by an NIHR Doctoral Fellowship and Google PhD Fellowship. HJM is supported by the NIHR Biomedical Research Centre at University College London.

## Supplementary materials

**Supplementary Material 1: Copy of study survey**

**Supplementary Material 2: Summary table of rank frequencies for each technique**

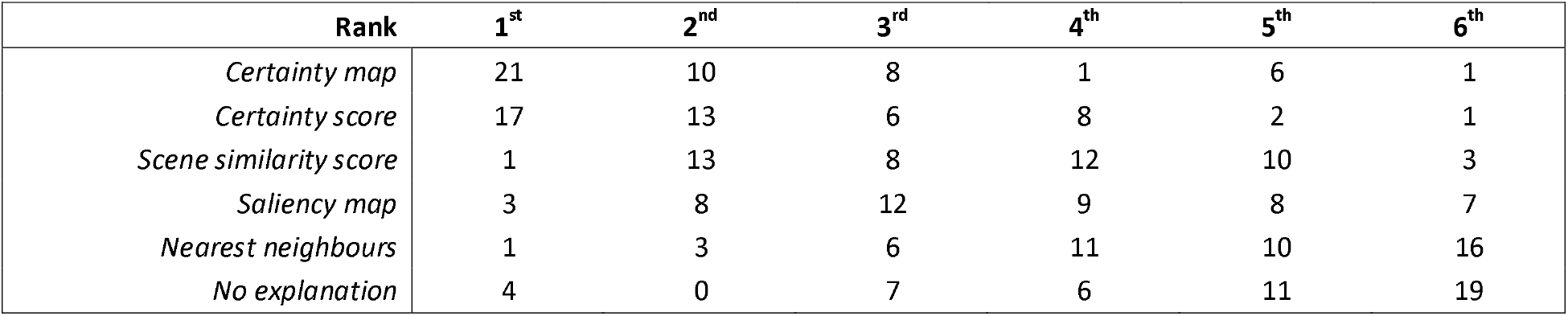

## Author Contribution Statement

- DZK - conception, design, analysis, first draft, revisions, final draft approval
- TJA - conception, design, analysis, first draft, revisions, final draft approval
- AW - conception, design, first draft, revisions, final draft approval
- RRH - conception, design, first draft, revisions, final draft approval
- SB - design, revisions, final draft approval
- PMC - analysis, revisions, final draft approval
- DS - conception, revisions, final draft approval
- MC - conception, revisions, final draft approval
- EC - conception, revisions, final draft approval
- AB - design, analysis, revisions, final draft approval
- HJM - conception, design, analysis, first draft, revisions, final draft approval
- CARES - data collection

## CARES Evaluation Group Author

- Mohammed Mustafa Adawi — Al-Zaitoun Specialized Hospital, Egypt

- Joao Paulo Almeida — Indiana University, USA

- Jensen Ang — National Neuroscience Institute, Singapore

- Anouk Borg — National Hospital for Neurology and Neurosurgery, London, UK

- Alexandros Boukas — Oxford University Hospitals NHS Foundation Trust, UK

- Aswin Chari — Institute of Child Health, University College London, UK

- Lekhaj C Daggubati — Washington Brain & Spine Institute, USA

- Neil L Dorward — National Hospital for Neurology and Neurosurgery, London, UK

- Samuel Emerson — University of Washington, USA

- Laurence Johann Glancz — Queen’s Medical Centre Nottingham, UK

- Paul Gwammache — Harvard University, USA

- John Gerrard Hanrahan — National Hospital for Neurology and Neurosurgery, London, UK

- Joachim Starup Hansen — National Hospital for Neurology and Neurosurgery, London, UK

- Lauren Harris — National Hospital for Neurology and Neurosurgery, London, UK

- Mark Hughes — Department of Clinical Neurosciences, Edinburgh, UK

- Abhiney Jain — National Hospital for Neurology and Neurosurgery, London, UK

- Adham M Khalafallah — University of Miami Neurosurgery Department, USA

- Taufiq Khan — Queen Elizabeth University Hospital, Birmingham, UK

- Muhammad Sohaib Khan — Lady Reading Hospital Peshawar, Pakistan

- Ramez Wadie Kirollos — National Neuroscience Institute, Singapore

- Angelos G Kolias — University of Cambridge, UK

- Jacek Kunicki — National Research Institute of Oncology, Poland

- Ruth Lau — Joan XXIII University Hospital, Spain

- Juan Carlos Fernandez Miranda— Stanford University, USA

- Ramesh Nair — Charing Cross Hospital, UK

- Nicola Newall — National Hospital for Neurology and Neurosurgery, London, UK

- Amr Nimer — Imperial College London, UK

- Fabio Massimo Oddi — University of Rome “Tor Vergata”, Italy

- Piero Andrea Oppido — Sapienza University of Rome, Italy

- Piotr Paździora — Medical University of Silesia in Katowice, Department of Neurosurgery, Poland

- Hafiz Muhammad Irfan Razzaq — Punjab Institute of Neurosciences, Lahore, Pakistan

- Bosnjak Roman — University Medical Centre Ljubljana, Slovenia

- Luis Alejandro Flores Salazar — Hospital Barros Luco Trudeau, Chile

- Olabisi Sanusi — Oregon Health & Science University, USA

- Thomas Santarius — Cambridge University, Cambridge University Hospitals, UK

- Sebastian Senger — Sozialstiftung Bamberg Department of Neurosurgery, Germany

- Jonathan Shapey — King’s College London, UK

- Nathan A Shlobin — Columbia University Irving Medical Center, USA

- Nicholas Wake Macmeikan Thomas — King’s College Hospital, UK

- Sebastian Miguel Toescu — National Hospital for Neurology and Neurosurgery, London, UK

- Georgios Tsermoulas — Queen Elizabeth Hospital in Birmingham, UK

- Juan Francisco Villalonga — LINT, Facultad de Medicina, Universidad Nacional deTucumán, Argentina

- Adam Williams — North Bristol NHS Trust, UK

- Nina Yoh — Columbia University Medical Center, USA

- Brett E Youngerman — Columbia University Medical Center, USA

- Gabriel Zada — University of Southern California, USA

## Notes

### Author Declarations

All methods were carried out in accordance with the principles of the Declaration of Helsinki and were approved by the University College London Ethics Committee (UCLIC-2024_009).

