## Supplementary Material 1 for "Exploring the Interpretability of AI Decision Support Systems for Surgical Anatomy Recognition"

### Exploring the Explainability of AI-Assistance for Anatomical Recognition in Endoscopic Pituitary Surgery

**Researchers:** Tobias J Adams, Danyal Z Khan, Anjana Wijekoon, Roxana Ramirez Herrera, on behalf of the Context Aware Augmented Reality for Endonasal Endoscopic Surgery (CARES) research team
**Principal Researchers:** Mr Hani J Marcus & Prof Ann Blandford

Thank you for considering participating in this study, which aims to gather your feedback on different ways of explaining decisions made by an artificial intelligence (AI) tool that predicts anatomical boundaries in endoscopic transsphenoidal surgery. Your feedback will inform the future development of this AI tool, allowing neurosurgeons to better utilise its decisions. In this study, explanations or supporting information are presented on a screen alongside the AI-predicted anatomical structure outlines. This simulates what may be seen when using the AI tool during surgery (e.g. to guide intra-operative decision-making) or after surgery on review of videos (e.g. for educational or training purposes).

For the best experience, this survey **should be completed on a desktop or laptop computer.** This survey will take **approximately 15 minutes** to be completed. We greatly appreciate your time and valuable insights.

As a participant in this study, **you have the option to be recognised as a collaborator author in our project under a collective group name**, called the “CARES Evaluation Group.” This form of collaborative authorship serves as an acknowledgement of your contribution without the responsibilities associated with primary authorship. Please note that opting into collaborative authorship is entirely voluntary, and you are free to participate in the survey without selecting this option. If you do choose to opt in, we will collect your name and affiliation for authorship purposes only. If you elect this option and provide your institution affiliation, you confirm that you have the authority to do so.

You are invited to participate if you:
- Are a neurosurgeon with at least one year’s experience in performing the endoscopic transsphenoidal approach
- Are able to communicate effectively in English
- Are able to give informed consent and do not consider yourself a vulnerable adult
- Do not have epilepsy or other conditions that may increase susceptibility to adverse reactions triggered by visual stimuli

No risks from taking part in this survey have been identified. In the unlikely event that participating causes you distress, you are free to withdraw and/or discuss your concerns with a listed researcher. If you have any concerns with the conduct of this study, please raise them first with Professor Ann Blandford. If your concerns are not addressed to your satisfaction, you may contact the Chair of the UCL Research Ethics Committee.

The results of this research project will be published in academic journals and presented at conferences. Summaries of the findings may also be shared with participants upon request. All published data will be anonymized to protect your identity. The controller for this project will be University College London (UCL). The UCL Data Protection Officer provides oversight of UCL activities involving the processing of personal data and can be contacted at. The only personal information retained will be a copy of your informed consent and your chosen contact details if you wish to be informed of the outcome of this study. These will be held securely and separately from the anonymised data that you provide for the study. Further information on how UCL uses participant information can be found here. The lawful basis that would be used to process your personal data will be performance of a task in the public interest.

#### Consent Form

**Title:** Exploring the Explainability of AI-Assistance for Anatomical Recognition in Endoscopic Pituitary Surgery
**Department:** UCL Interaction Centre (UCLIC), Department of Computer Science
**Researchers:** Tobias J Adams, Danyal Z Khan, Anjana Wijekoon, Roxana Ramirez Herrera
**Principal Researchers:** Hani J Marcus & Ann Blandford
**UCL Data Protection Officer:** Alexandra Potts
**Project ID number:** UCLIC-2024_009 (approved by the UCLIC Research Ethics Committee)

Please complete this consent form by selecting agree after reading the statements below.

**Statement 1**
I confirm that I satisfy all of the inclusion criteria:
- I am a neurosurgeon with at least one year's experience in performing the endoscopic transsphenoidal approach
- I am able to communicate effectively in English
- I am able to give informed consent and do not consider myself a vulnerable adult
- I do not have epilepsy or other conditions that may increase susceptibility to adverse reactions triggered by visual stimuli

**Statement 2**
I confirm:
- I understand all personal information will remain confidential and every effort will be made to protect my identity. However, if I choose to participate in collaborative authorship, absolute confidentiality cannot be guaranteed, as my name and affiliation will be included in a group authorship acknowledgment. This identifiable information will be securely stored separately from my survey responses and accessed only by authorised research team members.
- I understand that data collected will be stored in an anonymised form wherever possible and that my survey responses will not be directly linked to any personal information I provide. If I choose to participate in collaborative authorship, my name and affiliation will be collected separately and stored securely, ensuring that survey responses cannot be associated with my identity. I acknowledge that, according to data protection legislation, 'public task' will be the lawful basis for processing this data.
- I understand that my information may be subject to review by responsible individuals from the University for monitoring and audit purposes.
- I understand that I will not benefit financially from this study or from any of its future outcomes.
- I agree that my anonymised research data may be used by others for future research. No one will be able to identify me when this data is shared.
- I am aware of who I should contact if I wish to lodge a complaint.
- I understand that the information I have submitted will be published as a report.

Please select a response to the statement below:

I understand that I have the option to be recognised as a collaborator author under a group name as part of this study. This collaborative authorship means that my name and affiliation will be included in a group acknowledgement in any publications arising from this study, but I would not be responsible for primary authorship tasks or accountable for the study findings. I am aware that this option involves providing identifying information, and that anonymity cannot be guaranteed if I choose to opt in. If you elect to name your institution affiliation, you are confirming that you have the authority to do so.

Please provide your full name.

Full name of participant:

Date:

Please provide your email address if you wish to receive a copy of the report.

Email address:

How old are you?

- 18-24 years old
- 25-34 years old
- 35-44 years old
- 45-54 years old
- 55-64 years old
- 65+ years old

What is your biological sex?

- Male
- Female
- Prefer not to say
- Prefer to self-describe

Please select your level of experience:

- ○ Neurosurgery Consultant or Attending
- ○ Neurosurgery Senior Fellow (post-training)
- ○ Neurosurgery Registrar or Resident
- ○ [ ] Other (please specify)

How many years of experience do you have performing the endoscopic transsphenoidal approach?

- ○ 1 to 5 years
- ○ 5 to 10 years
- ○ 10 to 20 years
- ○ Over 20 years

In which areas of your practice do you use AI-assisted technologies? (Select all that apply)

- ☐ Academic research
- ☐ Administrative tasks (including medical record management)
- ☐ Clinical decision-making
- ☐ Disease diagnosis
- ☐ Medical imaging interpretation
- ☐ Robot-assisted surgery
- ☐ None
- ☐ [ ] Other (please specify below)

How often do you interact with AI-assisted technology in your practice?

- ○ a. Every day
- ○ b. Every week
- ○ c. At least once per month
- ○ d. At least once per year
- ○ e. Never

**Experimental Section 1**

### Explainable AI Methods for Anatomical Recognition in Endoscopic Pituitary Surgery

All of the anatomical outlines seen in this study are generated by an AI tool developed by our group, which has been trained on ~650 intraoperative images from endoscopic pituitary adenoma surgery. It uses a neural network to divide the intraoperative image into groups of pixels and analyse their characteristics - generating structure outlines.

You will now be shown intraoperative images immediately pre-sellotomy, with AI-generated outlines of key anatomical structures. The same AI-derived structure overlay is presented in each scenario, **however, the accompanying explanatory technique will differ.** These techniques seek to convey different levels of information about the AI's prediction, such as its confidence or its rationale.

For each different explanatory technique, you will be asked a series of questions to evaluate how well you think it explains or supports the AI's anatomical structure prediction.

**Certainty score**

#### Technique: Certainty score

This certainty score quantifies the AI model's certainty in identifying the area of each anatomical structure. The score represents the average pixel-wise certainty across the entire structure as a percentage. This visualisation is intended to allow for easy comparison of scores across multiple structures. It aims to help pinpoint which anatomical structures the AI model is certain about and highlight those that may require further evaluation or refinement. If the figure below is too small to view, please try the zoom function on your browser or view the figure on this link.

Structure outline legend: Green = Sella, Red = Carotid Protuberances, Pink = Optic Protuberances, Yellow = Clival Recess.

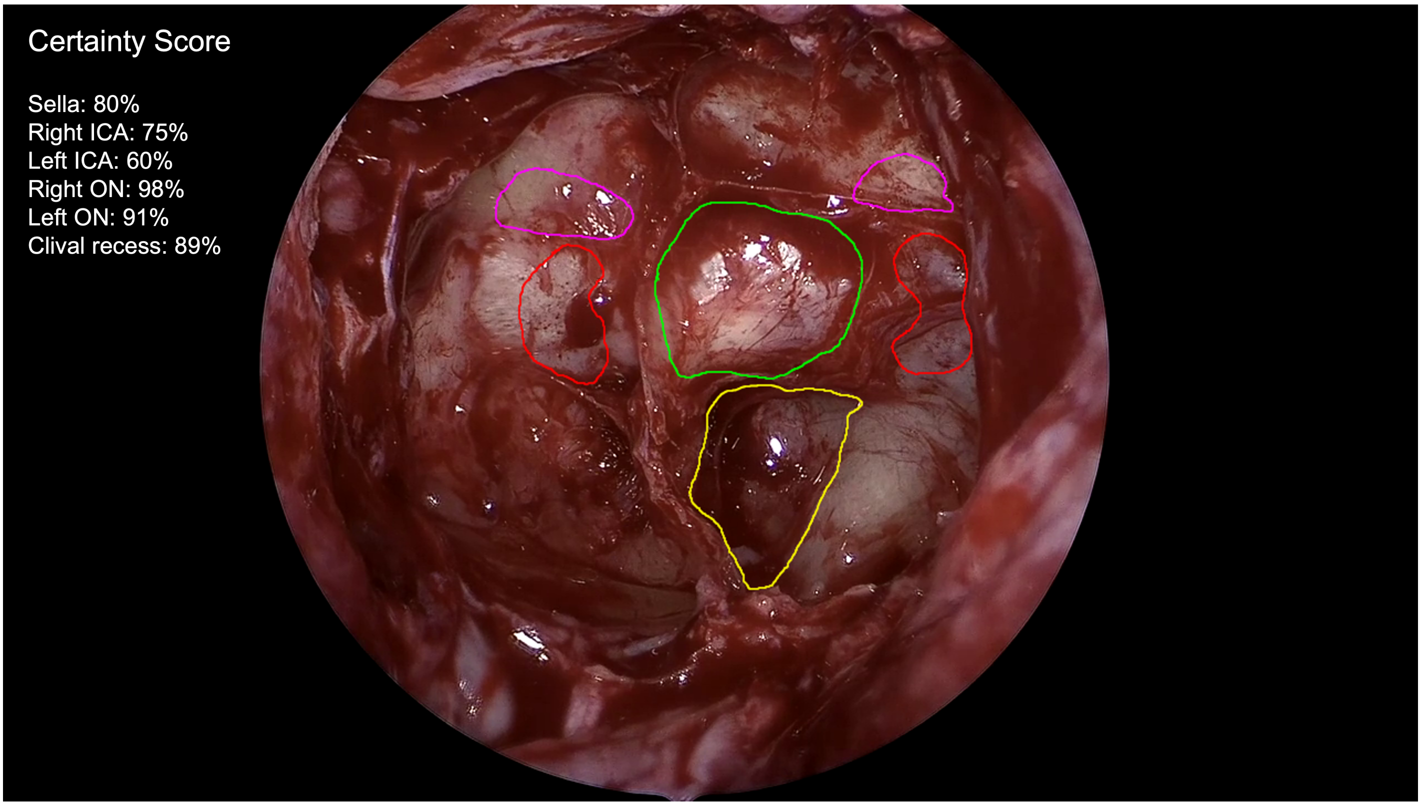


I'm neutral

| | I agree strongly | I agree somewhat | about it | I disagree somewhat | disagree strongly |
|---|---|---|---|---|---|
| This explanatory techique is **satisfying.** | ○ | ○ | ○ | ○ | ○ |
| This explanatory techique shows me how **accurate** the AI prediction is. | ○ | ○ | ○ | ○ | ○ |
| This explanatory techique has **sufficient detail.** | ○ | ○ | ○ | ○ | ○ |
| Using this explanatory techique, I **understand** how the AI works. | ○ | ○ | ○ | ○ | ○ |
| This explanatory techique is **useful to my surgical or educational goals.** | ○ | ○ | ○ | ○ | ○ |

Do you think this explanatory technique improves how clinically useful (e.g. for teaching or decision support) the AI outline overlay is? Please explain your rationale.

Optional Feedback

**Certainty map**

#### Technique: Certainty map

This certainty map (left) highlights the AI model's certainty in identifying the area for each anatomical structure. **Dark blue** colour indicates strong certainty, while **yellow-green** suggests areas where the AI model is less certain. By combining certainty maps for all anatomical structures into a single visualization, this aims to help compare AI model certainty across multiple structures at once. This visualisation also aims to help you to identify where anatomical structure precision might need further evaluation or refinement. If the figure below is too small to view, please try the zoom function on your browser or view the figure on this link.

Structure outline legend: Green = Sella, Red = Carotid Protuberances, Pink = Optic Protuberances, Yellow = Clival Recess.

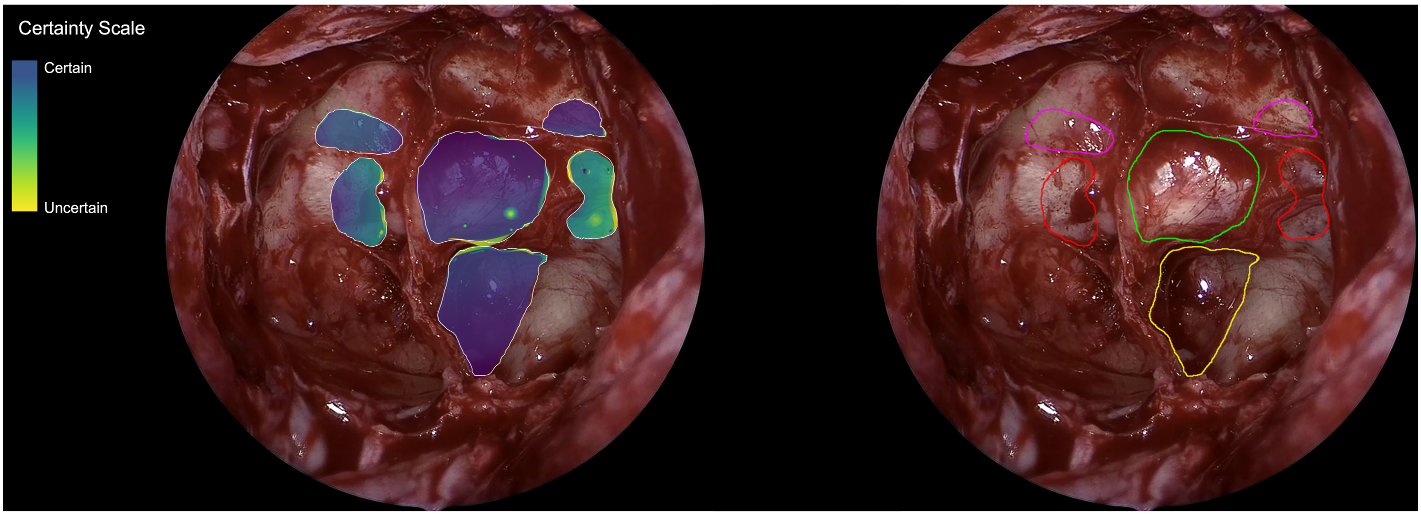


| | I agree strongly | I agree somewhat | I'm neutral about it | I disagree somewhat | I disagree strongly |
|---|---|---|---|---|---|
| This explanatory techique is **satisfying.** | ○ | ○ | ○ | ○ | ○ |
| This explanatory techique shows me how **accurate** the AI prediction is. | ○ | ○ | ○ | ○ | ○ |
| This explanatory techique has **sufficient detail.** | ○ | ○ | ○ | ○ | ○ |
| Using this explanatory techique, I **understand** how the AI works. | ○ | ○ | ○ | ○ | ○ |
| This explanatory techique is **useful to my surgical or educational goals.** | ○ | ○ | ○ | ○ | ○ |

Do you think this explanatory technique improves how clinically useful (e.g. for teaching or decision support) the AI outline overlay is? Please explain your rationale.

Optional Feedback

**Saliency map**

### Technique: Saliency map

This saliency map (left) highlights the regions in the image that the AI model considers most important for identifying anatomical structures. **Dark blue** areas indicate high saliency (where the model focuses most of its attention), while **green-turquoise** areas represent lower saliency (these contribute less to the model's predictions). By combining saliency maps for all anatomical structures into a single visualization, this aims to help compare the model's focus across multiple structures. This visualization aims to help you assess the consistency of the model's attention and identify areas where its focus might need further refinement. If the figure below is too small to view, please try the zoom function on your browser or view the figure on this link.

Structure outline legend: Green = Sella, Red = Carotid Protuberances, Pink = Optic Protuberances, Yellow = Clival Recess.

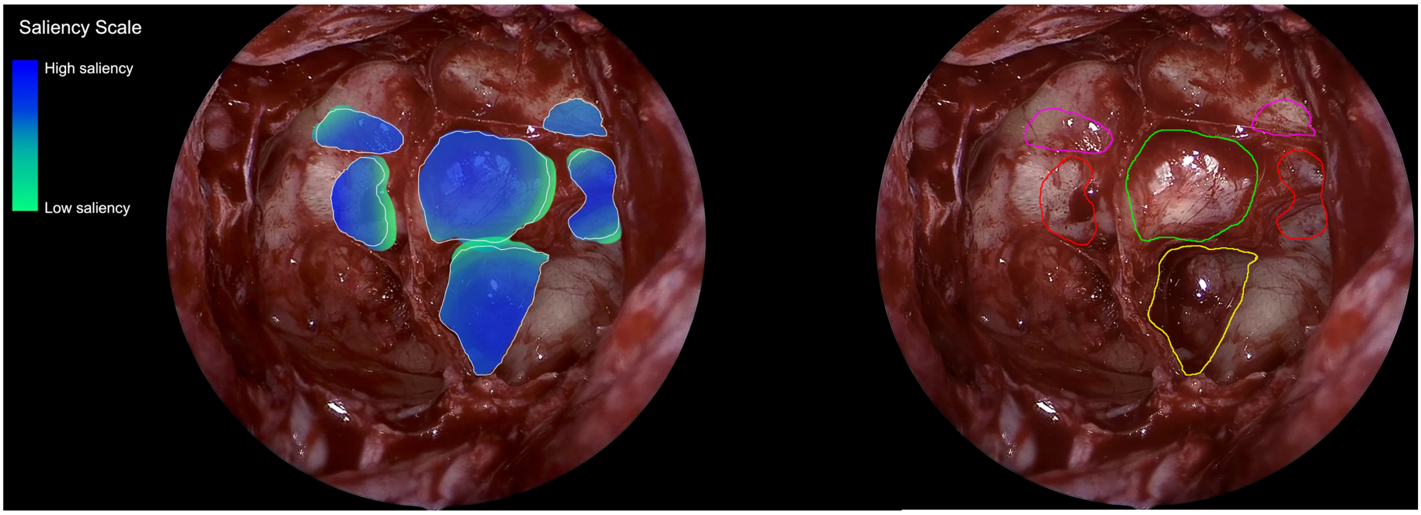


| | I agree strongly | I agree somewhat | I'm neutral about it | I disagree somewhat | I disagree strongly |
|---|---|---|---|---|---|
| This explanatory techique is **satisfying.** | ○ | ○ | ○ | ○ | ○ |
| This explanatory techique shows me how **accurate** the AI prediction is. | ○ | ○ | ○ | ○ | ○ |
| This explanatory techique has **sufficient detail.** | ○ | ○ | ○ | ○ | ○ |
| Using this explanatory techique, I **understand** how the AI works. | ○ | ○ | ○ | ○ | ○ |
| This explanatory techique is **useful to my surgical or educational goals.** | ○ | ○ | ○ | ○ | ○ |

Do you think this explanatory technique improves how clinically useful (e.g. for teaching or decision support) the AI outline overlay is? Please explain your rationale.

Optional Feedback

**Scene similarity score**

#### Technique: Scene similarity score

This scene similarity score shows how closely the current surgical scene matches the examples the AI model was trained on. A higher percentage suggests the model is working within its “comfort zone,” making it more likely that its predictions are accurate. In other words, when the image looks a lot like cases the AI already knows, it can apply its learned expertise more reliably. This score aims to give you a quick idea of how much confidence you might place in the model’s outputs. If the figure below is too small to view, please try the zoom function on your browser or view the figure on this link.

Structure outline legend: Green = Sella, Red = Carotid Protuberances, Pink = Optic Protuberances, Yellow = Clival Recess.

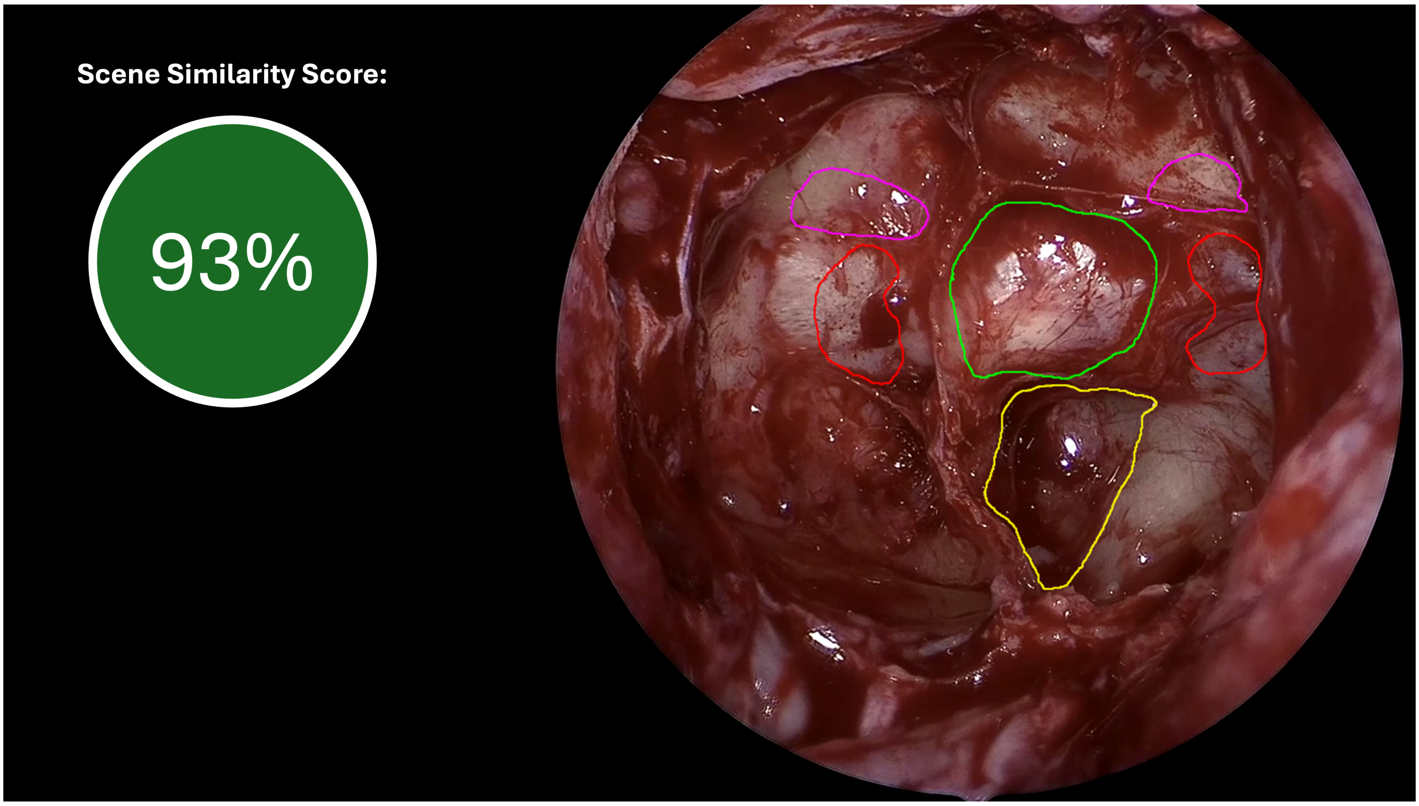

| | I agree strongly | I agree somewhat | I'm neutral about it | I disagree somewhat | I disagree strongly |
|---|---|---|---|---|---|
| This explanatory techique is **satisfying.** | ○ | ○ | ○ | ○ | ○ |
| This explanatory techique shows me how **accurate** the AI prediction is. | ○ | ○ | ○ | ○ | ○ |
| This explanatory techique has **sufficient detail.** | ○ | ○ | ○ | ○ | ○ |
| Using this explanatory techique, I **understand** how the AI works. | ○ | ○ | ○ | ○ | ○ |
| This explanatory techique is **useful to my surgical or educational goals.** | ○ | ○ | ○ | ○ | ○ |

Do you think this explanatory technique improves how clinically useful (e.g. for teaching or decision support) the AI outline overlay is? Please explain your rationale.

Optional Feedback

**Nearest neighbours**

#### Technique: Scene similarity (nearest neighbours)

This technique shows how similar the current image is to examples the AI was trained on and provides two "nearest neighbour" images for comparison. Each neighbour image, along with its similarity score, aims to help you understand how closely the current anatomy matches previously seen cases. By showing you what the model considers most similar, this aims to help you judge why it's making certain segmentation decisions. In essence, these reference images aim to serve as a window into the model's training experience, offering context for its current output. If the figure below is too small to view, please try the zoom function on your browser or view the figure on this link.

Structure outline legend: Green = Sella, Red = Carotid Protuberances, Pink = Optic Protuberances, Yellow =

Clival Recess.

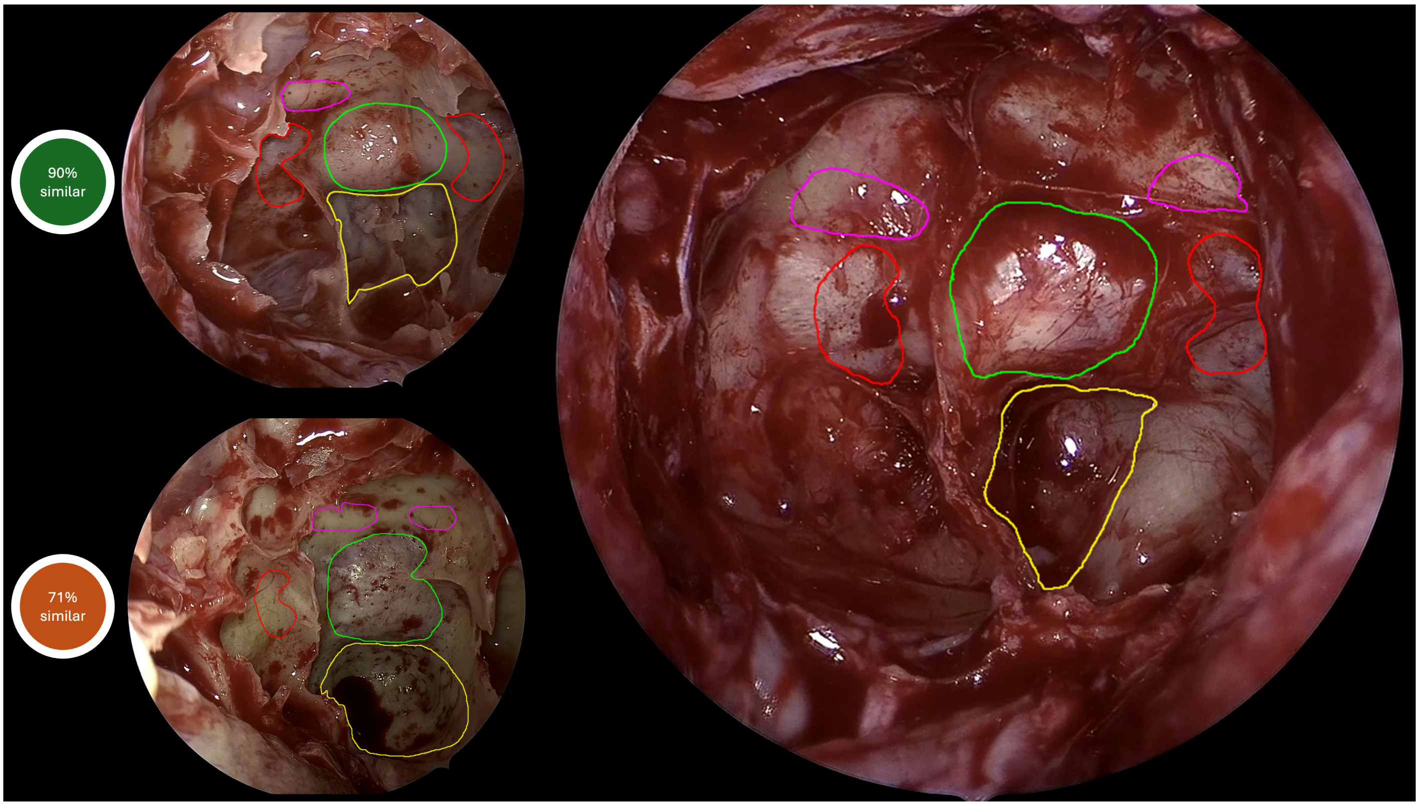


| | I agree strongly | I agree somewhat | I'm neutral about it | I disagree somewhat | I disagree strongly |
|---|---|---|---|---|---|
| This explanatory techique is **satisfying.** | ○ | ○ | ○ | ○ | ○ |
| This explanatory techique shows me how **accurate** the AI prediction is. | ○ | ○ | ○ | ○ | ○ |
| This explanatory techique has **sufficient detail.** | ○ | ○ | ○ | ○ | ○ |
| Using this explanatory techique, I **understand** how the AI works. | ○ | ○ | ○ | ○ | ○ |
| This explanatory techique is **useful to my surgical or educational goals.** | ○ | ○ | ○ | ○ | ○ |

Do you think this explanatory technique improves how clinically useful (e.g. for teaching or decision support) the AI outline overlay is? Please explain your rationale.

Optional Feedback

#### Ranking Question

### Overview and Ranking of All Explanation and Supporting Information Formats

You will now be shown all of the explanatory techniques previously displayed in the survey at once. You will be asked to rank them via dragging and dropping the options below, ordering them **from best (1/6) to worst (6/6)**. If the figure below is too small to view, please try the zoom function on your browser or view the figure on this link. There is also a free text box into which you may provide feedback or suggestions on the explanation techniques shown in this survey (optional).

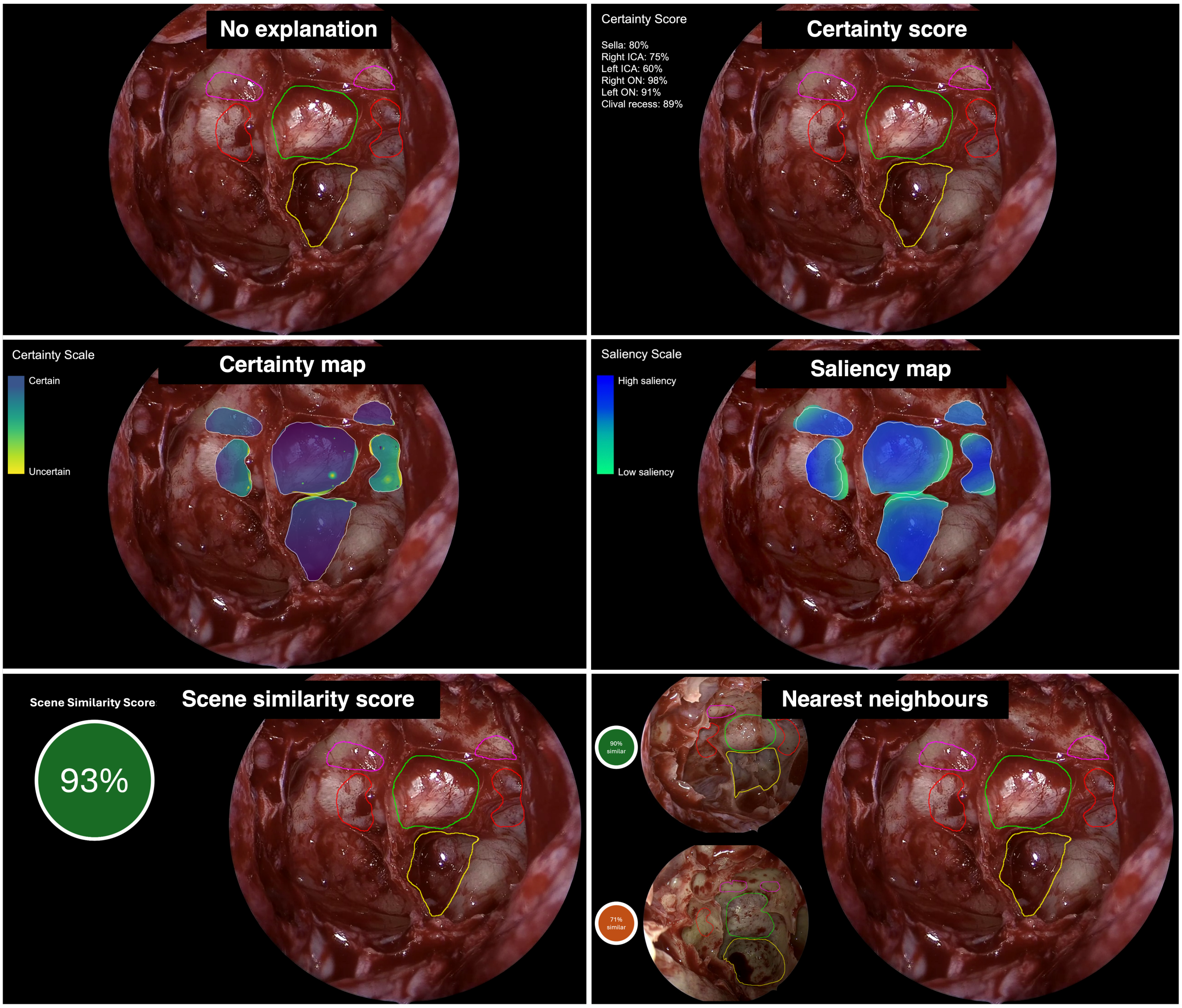

No explanation

Certainty score

Certainty map

Saliency map

Scene similarity score

Nearest neighbours

Optional Feedback

Powered by Qualtrics
